# Central Malaysia’s Silent Epidemic: Predictive Insights into Diabetic Foot Amputation

**DOI:** 10.1101/2025.10.12.25337839

**Authors:** Sanjiv Rampal, Claudia A Lopes, Parichehr Hadi, Vasantha Kumari Neela

## Abstract

Diabetic foot infection (DFI) is a prevalent and severe complication of diabetes mellitus, often leading to lower extremity amputations. This study uses electronic medical records from a tertiary care hospital in Central Malaysia to investigate the predictive factors associated with the type of amputation (major vs minor) in patients with DFI, once the amputation is decided. Methods: This retrospective cohort study analysed data from 434 patients admitted with DFI between January 2010 and December 2019. The information extracted from medical records included demographics, clinical characteristics (inflammatory markers and clinical complications), and amputation details. Binary logistic regression was performed to identify significant major vs minor predictors of amputation. Results: Most amputations were minor (70.7%), with Wagner grade 4 being the most common presentation. Elderly patients and higher Wagner scores are predictors of major (vs minor) amputation. Conclusion: This study underscores the critical role of age and Wagner classification for risk of major (vs minor) amputation among DFI patients. Identifying these underlying risk factors is crucial for developing targeted interventions, improving patient outcomes, and enhancing the quality of care for DFI patients.

## Introduction

Diabetes mellitus (DM) is one of the fastest-growing global health emergencies of the 21^st^ century, with its prevalence increasing at an alarming rate globally (45% increase in 5-years) [1]. Diabetes is a chronic disease that occurs either when the pancreas does not produce enough insulin or when the body cannot effectively use the insulin it produces. Hyperglycaemia, characterized by elevated levels of blood glucose (or blood sugar) is a common effect of uncontrolled diabetes and over which over time leads to serious damage to the heart, blood vessels, eyes, kidneys and nerves [45]. According to the International Diabetes Federation (IDF), approximately 589 million adults were living with diabetes in 2024, with three quarters of which living in low- and middle-income countries (LMICs). This figure is projected to rise to 853 million by 2050 [1]. The economic burden of diabetes is staggering, with 11.9% of global healthcare expenditure spent on diabetes (exceeding USD 1 trillion in 2024) [1].

The Western Pacific region has the highest number of people living with diabetes, accounting for 37% (215 million) of the total number of patients [1]. In this region, Malaysia has one of the highest diabetes prevalence rates, with 15.6% of adults affected — 1 in 6 adults [2]. An increasing trend in the overall prevalence of diabetes was seen from 2011 (11.2%), to 2015 (13.4%), and 2023 (15.6%), with a peak in 2019 (18.3%) [2]. One of the reasons is lifestyle as in Malaysia, 54.4% of adults are overweight (32.6%) and obese (21.8%) in 2023 and 84% of adults are not active in sports, fitness or leisure activities [2].

One of the most debilitating complications of diabetes is diabetic foot infection (DFI), which frequently leads to ulceration, gangrene, and ultimately, amputation [32]. Diabetic foot ulcers are the most serious and common consequence of diabetic mellitus. Studies suggest that 15-25% of patients develop a foot ulcer in their lifetime and up to 70% of these ulcers may become infected [3]. Amputation risk can be increased by factors such as age, smoking, abnormalities of the foot, poor glycemic control, ulcer size, hypertension, white blood cell count, and anomalies in lipids [4]. The risk of amputation among diabetic patients varies significantly across countries due to differences in healthcare infrastructure, glycemic control, and preventive measures. In high-income countries, amputation rates have declined due to multidisciplinary foot care teams and early intervention programs [5]. But in LMICs, amputation rates remain alarmingly high, primarily due to late presentation, limited access to specialized care, and poor glycemic control [6].

In Malaysia, diabetic foot complications account for nearly 70% of non-traumatic lower limb amputations, with major amputations (above the knee) being associated with higher mortality rates and reduced quality of life [7]. Minor amputations play a crucial role in limb preservation by removing nonviable tissue early, potentially preventing progression to more extensive amputations such as below-knee or above-knee procedures. A 10-year retrospective study in Hospital Kuala Lumpur reported that major amputations were more common among older patients (≥65 years) and those with peripheral arterial disease [8].

A study conducted in a Hospital in Pahang, Malaysia [45] involving patients with minor amputation (above ankle) identified poor compliance with diabetic treatment, King’s classification stage 5, low ankle-brachial systolic index measurements, presence of sensory neuropathy, elevated serum C-reactive protein, and elevated serum creatinine levels as significant predictive factors (p < 0.05). A previous study using the same sample as our study [44] analysed the distribution of microorganisms and prophylactic antibiotics. The results showed that among the Gram-negative bacteria, *Pseudomonas aeruginosa, Proteus* spp., and *Proteus mirabilis* were the most prevalent. Among the Gram-positive bacteria, the most identified organisms were *Staphylococcus aureus, Streptococcus agalactiae*, and methicillin-resistant *Staphylococcus aureus* (MRSA). The most frequently prescribed empirical antibiotics were ampicillin/sulbactam, followed by ciprofloxacin and ceftazidime. For therapeutic purposes, the most commonly used antibiotics were ampicillin/sulbactam, ciprofloxacin, and cefuroxime. Malaysia provides guidelines for management diabetic foot but lacks a standardized national diabetic foot screening program, leading to delayed referrals and higher amputation rates [9].

Some studies [10, 11, 12, 46] have proposed risk stratification models to predict amputation such as the SINBAD score (Site, Ischemia, Neuropathy, Bacterial Infection, Depth) validated in European, Asia and Africa cohorts (75% amputation risk for scores equal or higher than 4); The University of Texas Diabetic Wound Classification that incorporates infection and ischemia, showing 90-97% accuracy in predicting amputations (Grade 3, Stage D); and the Wagner score that considers depth of the wound and gangrene with 85% amputation risks for higher scores (4-5).

Implementing predictive models based on local data could significantly reduce preventable amputations. Therefore, identifying the risk factors for early management can reduce the amputation frequency among diabetic patients. This study aims to identify the predictive factors linked to type of amputation in patients who underwent amputations due to DFI, after the amputation was decided. The analysis predicts whether the amputation will be major vs minor based on a set of variables (sociodemographics, inflammation markers and comorbidities) retrieved from the records of patients from a Malaysian tertiary care hospital over a ten-year period, building on the evidence from previous studies.

## Material and methods

### Patient characteristics and clinical materials

This retrospective cohort study examined patients’ electronic medical records (EMR) of 434 patients with DFI who had their feet amputated at a tertiary care hospital (Hospital Ampang) in Central Malaysia between January 2010 and December 2019. The data was accessed between 8^th^ January 2024 and 31 May 2024. The EMR system included patients’ medical charts and clinical details such as medical history, medication, lab results, and doctor’s notes. The names and ID of patients were replaced with reference IDs prior to the analysis. The entries were made by healthcare providers as part of their clinical workflow for routine care. Therefore, the dataset is complete and accurate, comprising all patients hospitalized for foot amputation with DFI as their primary diagnosis. Pregnant women and those admitted for other foot infections, such as dry foot gangrene and dermatitis, were excluded from this study.

### Procedure selection for patients and variables

All patients initially received empirical antibiotic therapy based on the clinical severity of their infection. Following this, wound cultures were performed. Once culture results became available, treatment plans were adjusted accordingly. This included selecting appropriate targeted antibiotics (therapeutic antibiotics) and, when necessary, proceeding with amputation based on both the identified pathogens and the severity of the infection. The Wagner score was applied according to the Universal Wagner Classification and diagnosed by the specialist in charge who was part of the orthopedic multidisciplinary team of around 15 specialists with different clinical experience.

The information gathered from patients’ records included ethnicity, age, gender, smoking habits, Wagner grade, inflammatory markers (ESR, WBC CRP), neuropathy, peripheral vascular disease (PVD), osteomyelitis, gangrene, and amputation type including major (above knee) and minor (at the level of knee and below) amputation. The data was recorded in Excel and analysed in R Studio. The data preparation, analysis and visualization code are published online [13].

### Statistical analysis

The distributions of all variables are presented and analysed using descriptive statistics, such as frequency analysis for categorical variables, mean, standard deviation (SD) for interval variables and median, and interquartile range (IQR) for ordinal or not normally distributed interval variables. Bivariate associations between each one of the risk factors and amputation risk were analysed using the Mann-Whitney U test for interval variables, the z-test to compare percentages between two groups and the chi-square test to establish associations between categorical variables. The risk of amputation was analysed through a binary logistic regression model, considering the risk factors with significant associations (p<.05) with the type of amputation. The odds ratio (OR) and respective confidence intervals (CI) were obtained for all variables in the model, and the significance was considered for interpretation.

## Results

### Sociodemographic Characteristics

The sociodemographic characteristics of DFI patients at Hospital Ampang are shown in Table 1. More than half of the patients (56%) are aged between 41 and 65 years, followed by those over 65 (39.2%), with 4.8% being between 18 and 40. Among the patients, 62.9% were male and 37.1% were female. Across all major ethnic groups in Malaysia (Malay, Indian, and Chinese), the Indian patients had the lowest representation at 15%, while Malay patients were the most represented at 61.8%. The Malaysian ethnicity demographics comprises approximately 69.9% Malays (Bumiputras), 22.8% Chinese, 6.6% Indians, and about 0.7% others. Additionally, the majority (61.2%) of the patients were found to be non-smokers. All demographic information was complete except for smoking status that contained 15% of missing values.

**Table 1.** Association between sociodemographic characteristics and type of amputation (Chi-square test for independence, column percentages).

| Variable | Level | Frequency | Minor, n (%) | Major, n (%) | p-value |
| --- | --- | --- | --- | --- | --- |
| Age Group | 18–40 years | 21 (4.8%) | 18 (5.9%) | 3 (2.4%) |  |
|  | 41–65 years | 243<br>(56.0%) | 184<br>(59.9%) | 59 (46.5%) |  |
|  | >65 years | 170<br>(39.2%) | 105<br>(34.2%) | 65 (51.2%) | .003** |
| Sex | Female | 161<br>(37.1%) | 113<br>(36.8%) | 48 (37.8%) |  |
|  | Male | 273<br>(62.9%) | 194<br>(63.2%) | 79 (62.2%) | .932 |
| Ethnicity | Chinese | 91 (21.0%) | 63 (20.5%) | 28 (22.0%) |  |
|  | Indian | 65 (15.0%) | 50 (16.3%) | 15 (11.8%) |  |
|  | Malay | 268<br>(61.8%) | 184<br>(59.9%) | 84 (66.1%) |  |
|  | Other | 10 (2.3%) | 10 (3.3%) | 0 (0.0%) | .112 |
| Smoking Status | Smoker | 98 (26.6%) | 78 (29.4%) | 20 (19.2%) |  |
|  | Non-Smoker | 226<br>(61.2%) | 159<br>(60.0%) | 67 (64.4%) |  |
|  | Ex-Smoker | 45 (12.2%) | 28 (10.6%) | 17 (16.3%) | .073 |

Note. n = sample size. Significance: \* $p < .05$ , \*\* $p < .01$ , \*\*\* $p < .001$ .

The breakdown of sociodemographic characteristics by type of amputation is also presented in Table 1. Most patients had minor amputations (n=307; 70.7%), while 29.3% (n:127) had major amputations. According to the chi-square test of independence, age group was found to be significantly associated with the type of amputation (*p* <.01). These findings indicate that major amputations were more common among elderly patients aged ≥65 (51.2%). In contrast, minor amputations were more common among patients aged 41–65 (59.9%).

### Indicators of Inflammation

Several markers could be used in measuring the inflammatory response in diabetic foot infection patients such as C-reactive protein (CRP), erythrocyte sedimentation rate (ESR), procalcitonin (PCT), and white blood cell count (WBC). Elevated levels of these markers indicate the presence and severity of infection, aiding in diagnosis and monitoring treatment response. Cytokines such as Interleukins (ILs) and Tumor Necrosis Factor-alpha (TNF-α) can be elevated in diabetic foot infections, however, they are not as commonly used as CRP (inflammation at an acute stage), and ESR (indicator of bone infections). Hence the studied hospital panel is restricted to WBC, CRP, and ESR.

The mean±SD of WBC is 17.68±7.54 x 10^3^/μL, with most patients having abnormal levels over 11 x 10^3^/μL (82.1%), with the percentage of missing values below 1%. The mean±SD for CRP is 170.33±109.97 mg/L, with all patients having an abnormal CRP value over 5 mg/L. However, this data was not available for 47.5% of the patients. Finally, the mean±SD for ESR is 96.98±28.18 mm/hr. This value was missing for 39.6% of patients.

Since the inflammation markers present a skewed distribution, the Mann-Whitney test was used to compare the mean difference between the two types of amputation (Table 2). The results showed that WBC, CRP, and ESR median levels differ significantly by type of amputation. Across all markers, patients with major amputations showed higher levels of these inflammation markers.

**Table 2.** Association between inflammation markers and amputation (Mann-Whitney U test).

| Markers | Minor, median (IQR) | Major, median (IQR) | p-value |
| --- | --- | --- | --- |
| WBC | $15.5 (9.22) \times 10^3/\mu\text{L}$ | $18 (9.15) \times 10^3/\mu\text{L}$ | <.001*** |
| CRP | 139 (172) mg/L | 191 (123) mg/L | .002** |
| ESR | 101 (28) mm/hr | 108 (29) mm/hr | .021* |

Note. IQR = Interquartile range. Significance: \* $p < .05$ , \*\* $p < .01$ , \*\*\* $p < .001$ .

### Complications from Diabetes

The duration of diabetes varies between 1 and 37 years with mean±SD of 10.95±8.1 years (the percentage of missing values is 37.1% of patients). The most frequent diabetic complications were gangrene (66.4%), osteomyelitis (64.2%), and peripheral neuropathy (25.3%) followed by peripheral vascular disease (13.8%). The test of association between amputation and complications from diabetes revealed that all complications were significantly associated with the type of amputation (Table 3).

**Table 3.** Association between complications from diabetes and amputation categories (Two proportion z-test, row percentages).

| Complications | Frequency (%) | Minor, n (%) | Major, n (%) | p-value |
| --- | --- | --- | --- | --- |
| Gangrene | 188 (66.4%) | 133 (70.7%) | 55 (29.3%) | <.001**<br>* |
| Peripheral Neuropathy | 110 (25.3%) | 87 (79.1%) | 23 (20.9%) | <.001**<br>* |
| Osteomyelitis | 185(64.2%) | 139 (75.1%) | 46 (24.9%) | <.001**<br>* |
| PVD | 60 (13.8%) | 36 (60%) | 24 (40%) | <.05* |

Note. n = sample size. Significance: \* $p < .05$ , \*\* $p < .01$ , \*\*\* $p < .001$ .

Based on the Wagner classification (Table 4), the patients’ distribution indicated that Wagner grades 4 and 5 are the most common (40.3% and 23.5%, respectively). The relationship between Wagner classification and amputation was examined using the chi-square test. The results in Table 4 indicate a significant relationship between Wagner classification and amputation (X^2^(4) =288.90, *p*<.001). Grades 1-4 often resulted in minor amputations, while major amputations were mainly performed in grade 5 (79.5% of major amputations).

**Table 4.** Association between Wagner grade and type of amputation (Two-proportion z-test, column percentages).

| Level | Frequency (%) | Minor, n (%) | Major, n (%) | z-statistic | p-value |
| --- | --- | --- | --- | --- | --- |
| Grade 1 | 9 (2.1%) | 8 (2.6%) | 1 (0.8%) | 3.30 | <.001**<br>* |
| Grade 2 | 77 (18%) | 67 (21.8%) | 10 (8.2%) | 9.19 | <.001**<br>* |
| Grade 3 | 66 (15.4%) | 64 (20.8%) | 2 (1.6%) | 10.79 | <.001**<br>* |
| Grade 4 | 173 (40.3%) | 161 (52.4%) | 12 (9.8%) | 16.02 | <.001**<br>* |
| Grade 5 | 105 (24.0%) | 6 (2.0%) | 97 (79.5%) | -12.68 | <.001**<br>* |

Note. n = sample size. Significance: \* $p < .05$ , \*\* $p < .01$ , \*\*\* $p < .001$ .

### Binary Logistic Model

We analysed the effect of the different variables on the odds of undergoing a minor vs major amputation through binary logistic regression results (Table 5) considering age, duration of illness, inflammatory markers, complications (osteomyelitis, peripheral neuropathy and peripheral vascular disease), and Wagner grade. The results revealed that age and Wagner score are the main predictors of minor vs major amputation.

**Table 5.**
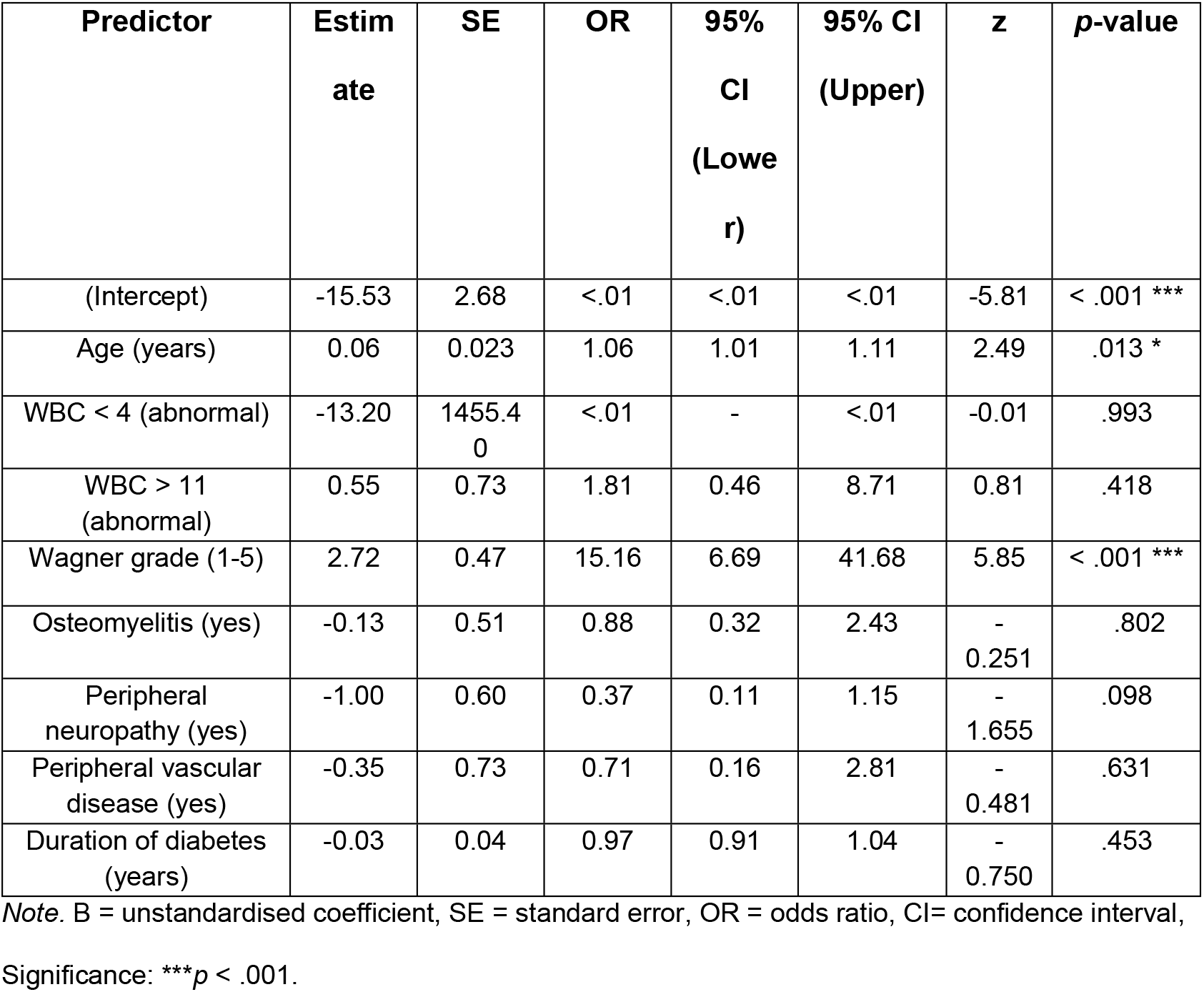
Binary logistic regression predicting odds of major (vs minor) amputation (AIC: 206.87).

**Table 6.**
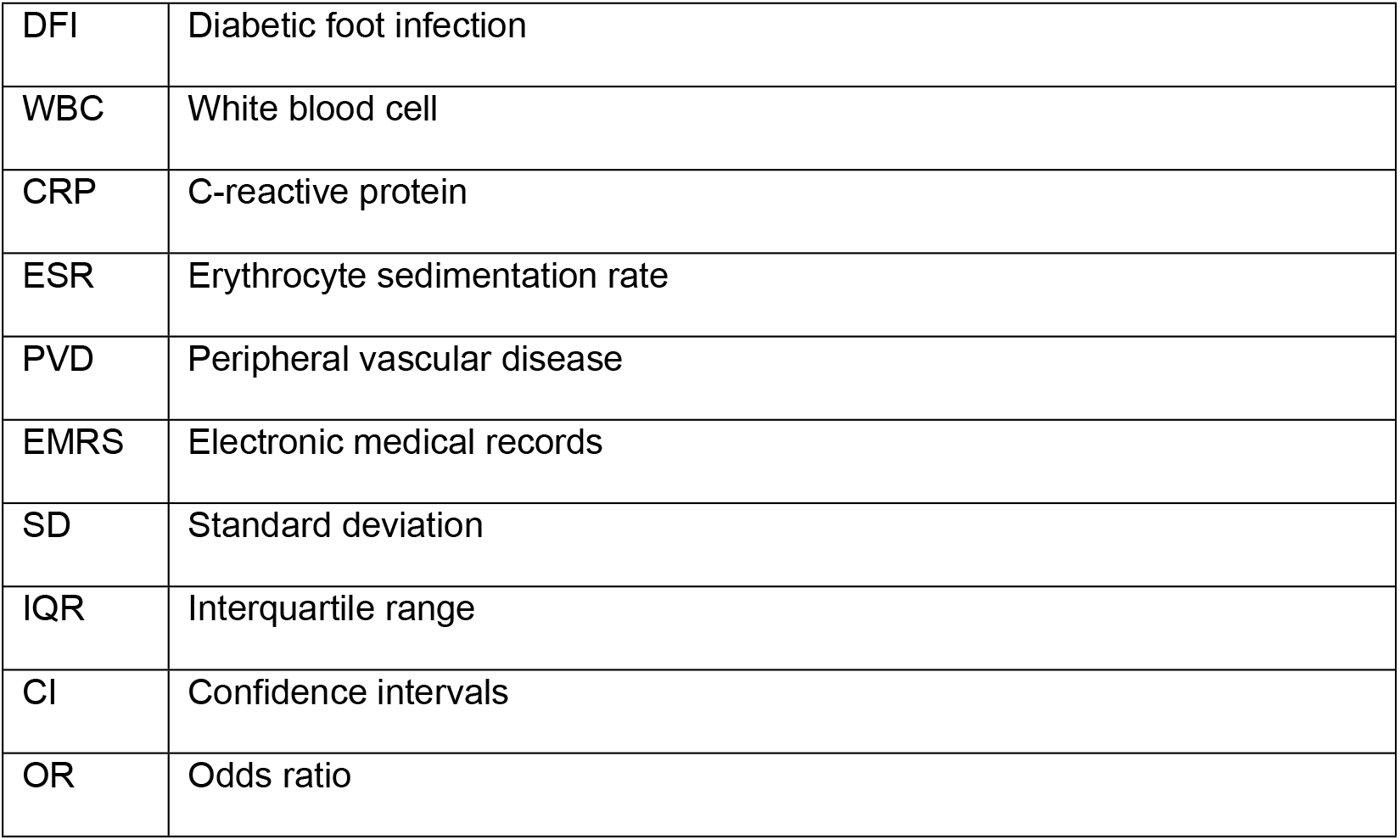
The following abbreviations are used in this manuscript:

|  |  |
| --- | --- |
| DFI | Diabetic foot infection |
| WBC | White blood cell |
| CRP | C-reactive protein |
| ESR | Erythrocyte sedimentation rate |
| PVD | Peripheral vascular disease |
| EMRS | Electronic medical records |
| SD | Standard deviation |
| IQR | Interquartile range |
| CI | Confidence intervals |
| OR | Odds ratio |

Only one inflammation marker was considered (WBC) because ESR and CRP had a high percentage of missing values (over 40%). Gangrene was also excluded from the model because it was moderately associated with Wagner score [X^2^(4) = 46.201, p-value < .001, Spearman rho = 0.26]. This result is expected as gangrene (partial or extensive) is one of the criteria for Wagner scores 4 and 5, respectively. In this study, 64.3% of the patients had Wagner grade 4 or 5. Wagner.

We tested multicollinearity through the variance inflation factor (VIF) and all the predictors showed an index value of no multicollinearity (values between 1.0 and 1.1). The McFadden’s Pseudo R^2^ = 0.636 (<and the Cragg-Uhler R^2^=0.893, denoting that the model has a strong explanatory power.

## Discussion

Diabetes raises the risk of foot problems, which can rapidly progress and, in the presence of co-morbidities, lead to amputation. In addition to being a medical problem, this condition significantly lowers patients’ quality of life and well-being. In this section, we present the main results of this study, discussing them in the light of other previous studies in Malaysia and other countries. It is worth noting that this is a single hospital-based study that needs confirmation in a multi-center study, although we triangulate these results with results from studies in two other hospitals in Malaysia [15, 18, 19].

In this study, 51.2% of major amputations were done in individuals over 65 and age was identified as predicting the risk of patients undergoing a major (vs minor) amputation. The most common age group for hospitalised DFI patients was 41-65 years. DFI was more common among male compared to female patients (62.9% and 37.1%, respectively). A higher infection rate was found among the Malays (61.8%), followed by the Chinese (21%) and the Indians (15%). As Malay ethnicity accounts for most of the Malaysian population (about 70% of population), diabetic foot problems are a prevalent problem. The different prevalence across ethnicities is related to lifestyles and obesity rates [25]. These demographic differences found in the sample were not significantly associated with the risks of major vs minor amputation.

In our study, WBC, CRP, ESR levels were significantly higher (p < 0.05) in DFI patients who later undergo major amputations compared to those who had minor amputations. Clinical complications such as gangrene, peripheral neuropathy, osteomyelitis, and peripheral vascular disease were also significantly associated with major amputation in the univariate analysis.

To further examine predictors of amputation severity, a binary logistic regression model was conducted including Wagner grade, WBC groups (normal and high/low abnormal), and diabetes-related complications (peripheral neuropathy, PVD, and osteomyelitis), while controlling for age and duration of illness. Among these variables, only Wagner grade and age emerged as significant independent predictors of major versus minor amputation (p < 0.0001) and WBC groups and diabetes-related complications did not remain significant in the multivariate model. This attenuation is related to patients with Wagner grade 5 having elevated WBC values, and diabetes-related complications tending to cluster in older patients and those with longer disease duration.

A study from Ethiopia also found that most DFI patients are aged 58–67 [14]. Similarly, a different study in Malaysia agrees with our findings, indicating that the mean age of DFI patients was 58.6 years [15]. A Korean study also revealed an association between age and amputation with the average age of amputation of 61.1 years [16]. Conversely, a study conducted in India found no association between the DFI patients’ age and the type of amputation (*p* = 0.566) [17].

Previous studies in Malaysia also revealed that DFI patients are predominantly male (59.2%) [18]; and that male DFI patients had a greater infection rate (61.0%) compared to females (39.0%) [19]. This could be attributed to the lifestyle of men who are more commonly involved in physical activities, working long hours, not able to visit clinics for the regular follow up due to their job nature which delays the recovery of ulcers and increases the chances of amputation. Though similarly to other studies [20,21] where gender and amputation were not associated, contradictory results emerged in studies that identified gender as a risk factor for amputation [22, 23,24].

In relation to ethnicity, our findings are consistent with other studies. For example, at Hospital Segamat (HS) in Johor, Malaysia [26] among 595 diabetic foot infection patients, the greater proportion were Malays, followed by Chinese, and Indians. Another study from Pahang showed that most patients were Malays, followed by Chinese and Indian [25]. Notwithstanding, it has also been reported that, in Malaysia, ethnicity plays a role in diabetes complications, with Indians having the highest risk of DFI-related amputations [39]. Though smoking does not show significant differences between the minor and major amputation groups, it is a notable risk factor for diabetic foot amputation [47].

Elevated WBC, CRP, and ESR indicate severe infection and systemic inflammation [27]. Several studies corroborate the bivariate associations found between inflammation markers and major (vs minor) amputation. A prospective investigation into risk variables for amputation in 379 DFI patients in Turkey revealed a strong and significant correlation (*p* <0.001) between amputation and ESR and CRP [28]. Similarly, studies in Singapore found that CRP >100 mg/L was a risk factor of limb loss [29]; and WBC was a significant risk factor of amputation in another study [30]. In a retrospective analysis (2016-2019) of 320 patients with DFI in a tertiary hospital in Malaysia (Hospital Serdang), elevated WBC >12,000/μL was associated with a 3-times increase; and CRP >80 mg/L was associated with 4.3-times increase in amputation risk [31].

PVD impairs wound healing due to reduced blood flow and neuropathy, allowing minor injuries to progress into severe infections, and increasing the risk of gangrene. A meta-analysis found that PVD increased amputation risk by 4.5 times, while neuropathy increased it by 3.2 times [32]. A study in Turkey found a correlation (*p* = 0.012) between neuropathy and amputation [40].

Localized gangrene is present in ulcers categorised as Wagner grades 4 and 5, which tend to be brought on by ischaemia and infection. Wagner classification has been positively associated with the risk of amputation [33, 34]. Similarly to our study, other studies in India [35], Thailand [36], and Korea [37] also revealed grades 4 and 5 were strongly linked with major amputations. This result also agrees with another study in Malaysia that reported that 80% of major amputations occurred in patients with Wagner grade 4 or 5 ulcers [38]. Other studies in India and Iran also revealed Wagner grades 4-5 and delayed presentation (over 30 days) worsened outcomes, while early surgical debridement reduced amputation [39, 40]. Thus, the association between amputation and grade 4 and 5 ulcers is not surprising.

When all factors are considered, some predictive factors appear more relevant. For example, a study with 586 hospitalized patients with moderate-to-severe DFIs in the US identified 5 predictors of amputation: PVD, wound necrosis, elevated CRP (>13mg/L), prior amputation and lack of oral diet (as a marker of severe illness) [27]. A study in Turkey with 250 patients with DFI found that the top risk factors for amputation were osteomyelitis, Wagner grade 4 and 5, PVD, and elevated inflammatory markers (CRP >80 mg/L and ESR >70 mm/h) [41].

One important difference between our study and the other studies cited is that ours distinguishes the risk of major vs minor amputation, once the amputation decision was made. Other studies are limited to identifying risk factors for amputation among patients with DFI, some of whom are not going to undergo amputation. An exception is a cohort study in Turkey with 186 DFI patients (2012-2015) which analysed risk factors and outcomes of minor vs major amputation. This study identified PVD and Wagner grade as major predictors of major vs minor amputations [42].

The economic condition of the patient was identified as an important factor in a study with 1850 patients from LMICs, including Malaysia, Pakistan, and Egypt [43]. The study showed that low-income patients were 3.2 times more likely to present with advanced infections (Wagner grade 4–5) compared to higher-income groups. The main reasons for delaying healthcare were related to the cost of care (52%), lack of transport (34%), and low health literacy (29%). The income level of the patient was not considered in our study and therefore discussed in the limitations.

One limitation of this study is that it included only DFI patients who underwent amputation, in order to assess the risk factors that distinguish minor versus major amputation. Patients with DFI who did not undergo amputation were not included. As a result, the findings cannot be generalized to the broader population of DFI patients at risk of amputation, but only to the subset in whom amputation was already indicated or performed. Additionally, this study relies on data from a single tertiary hospital in Central Malaysia, which inherently restricts the generalizability of our findings. This single-center approach, coupled with the unique characteristics of Malaysia’s dual public and private healthcare system, where surgical theatre access is predominantly in state capital hospitals and specialist care referral pathways vary considerably between rural and urban peripheries, likely contributed to increased morbidity in the study cohort.

Beyond this, crucial confounders, including HbA1c levels, patient income level, and delay in presentation, could not be comprehensively included in our study. This was due to a combination of factors prevalent in this setting: poor standardization of investigation and care protocols across referring facilities, significant interpersonal differences in care delivery, and challenges in accessing detailed income level information for individual patients due to stringent Personal Data Protection Act (PDPA) practices. Furthermore, HbA1c levels, a crucial indicator of glycemic control, were unfortunately investigated in less than 25% of patients, largely owing to the perceived high cost and a prevailing preference for the use of fasting blood sugar measurements in routine practice. These unaddressed confounders, particularly the impact of the multi-tiered healthcare system on patient presentation severity and the limitations in comprehensive data capture for key diabetic parameters and socioeconomic factors, warrant a more detailed reflection on how they may have affected the observed outcomes and limit the broader applicability of our conclusions to other settings.

## Conclusion

This retrospective cohort study identified Wagner grade 4–5 ulcers and older age (≥65 years) as the main predictors that distinguish between minor vs major amputation among DFI patients in a tertiary care hospital in Malaysia. Secondary factors included elevated inflammatory markers (WBC, CRP, and ESR) and complications such as gangrene, peripheral neuropathy, osteomyelitis, and peripheral vascular disease. To improve outcomes globally, it is recommended that healthcare systems prioritize early detection and aggressive management of DFI, especially in high-risk patients presenting with advanced Wagner grades, older age, peripheral vascular disease, peripheral neuropathy, or elevated inflammatory markers. Furthermore, allocating adequate resources for DFI patient care and fostering international collaborative research to uncover additional risk factors and innovative treatment strategies are essential for global amputation prevention efforts.

## Data Availability

All relevant data are within the manuscript and its Supporting Information files

https://rpubs.com/cimdal2/DFI

## Abbreviations

## Acknowledgments

The authors thank all members in the hospital who helped with the data collection. This study was not supported by any research grant.

## Ethical approval

The study was carried out according to the guidelines of the Declaration of Helsinki, and approved by Medical Research and ethics committee, Ministry of Health Malaysia, NMRR ID-22-00250-PB8 (IIR),date: 29 April 2022.

## Informed Consent Statement

Patient consent was not required, as the study was based on retrospective data and there was no direct contact with the individuals involved.

